# Health shocks and changes in life purpose: Understanding the link between purpose and longevity

**DOI:** 10.1101/2022.03.13.22272313

**Authors:** Richard Sias, H. J. Turtle

## Abstract

**Background:** The negative correlation between life purpose levels and subsequent morbidity and mortality is interpreted as evidence that a higher sense of life purpose causes healthier and longer lives. Causation, however, could run the other direction as a decline in health is, by definition, associated with greater morbidity and mortality risk and may also cause a decline in life purpose. We examine the relation between objective measures of changes in health and changes in purpose to better understand the causal mechanisms linking purpose to health and mortality.

**Methods:** Prospective cohort sample of 12 745 individuals aged 50 and older who were eligible to participate in the 2006, 2010, or 2014 Health and Retirement Study Psychosocial and Lifestyle questionnaire. The final sample consists of 15 034 observations measured over three four-year periods from 5 147 individuals. Controlling for standard covariates, we examined the relation between changes in purpose and 14 contemporaneous and subsequent objectively measured changes in health—lung function, grip strength, walking speed, balance, and physician diagnoses of hypertension, diabetes, cancer, lung disease, heart condition, stroke, psychiatric problem, arthritis, dementia, and Alzheimer’s disease.

**Findings:** There is strong evidence that negative health shocks cause a decline in life purpose as individuals who suffer a negative health shock experience a statistically meaningful contemporaneous decline in life purpose for 12 of the 14 changes in health metrics. In contrast, there is relatively weak evidence that a decline in purpose contributes to a deterioration of future health.

**Interpretation:** Much of the relation between life purpose levels and mortality risk arises from reverse causation—a decline in health causes both increased mortality risk and lower life purpose. There is little evidence that life purpose interventions would alter future morbidity or mortality.

**Funding:** None.

**Research in context:** *Evidence before this study:* We searched PubMed and Google Scholar with no language or date restriction for the term “life purpose” and found four comprehensive reviews of the life purpose or psychological well-being (which included life purpose in the set of psychological well-being metrics) literatures in the last three years and a 2016 meta-analysis of the relation between life purpose and mortality. Although acknowledging the possibility that reverse causation plays a role in linking life purpose levels to subsequent morbidity and mortality, the prevalent view appears to be that even when controlling for current health levels, higher life purpose causes behavioral, biological, or stress buffering changes that, in turn, cause lower future morbidity and mortality.

*Added value of this study:* By focusing on changes in health, changes in life purpose, and a longer horizon, we find strong evidence that changes in health cause changes in life purpose, but, contrary to the conclusions of most previous work, there is little evidence changes in life purpose cause changes in behavior, biology, or stress-buffering that, in turn, cause changes in future health.

*Implications of all the available evidence:* Although life purpose intervention—either at the provider level or in public policy—may have benefits, there is little evidence to suggest it will cause greater longevity or lower future illness.

## Introduction

Previous work finds that higher levels of purpose in life are associated with lower future illness and mortality and proposes that life purpose intervention may offer a promising low-cost opportunity to improve health and longevity.^1–16^ A 2016 meta-analysis, for example, reports that higher life purpose is associated with a relative risk of 0·83 for both all-cause mortality (*p*<0·001) and cardiovascular events (*p*<0·001).^17^ Although the evidence is promising, causation is less clear as the link between purpose and mortality may arise because: (1) low purpose causes decreased health and increased mortality risk, (2) a decline in health causes increased mortality risk and lower purpose (i.e., reverse causation), or (3) both paths contribute (although not necessarily equally) to the relation between purpose and subsequent morbidity and mortality. In short, if a decline in health causes lower purpose, then reverse causation must play some role in explaining the purpose-mortality relation (unless one is able to perfectly control for health) and, although life purpose will be correlated with subsequent mortality risk, low purpose might not be the only, or even primary, causal factor.

Although acknowledging the potential for reverse causation, most work focuses on testing whether life purpose levels are associated with subsequent mortality, behaviors, or illness. Because this work often includes measures of current health or excludes individuals with negative outcomes in the first year, the consensus view holds that reverse causality is unlikely to play a substantial role in driving the purpose-mortality (including CVD) relation, e.g., “*…a majority of studies evaluating the purpose–CVD connection already incorporate some methodological strategies (e*.*g*., *adjusting for a range of cardiovascular risk factors at baseline) which can partially address potential concerns about reverse causality. Thus, it seems unlikely that even if future studies consistently account for all of these factors, they will overturn findings from existing studies*.”^18^

We take a new approach to examining the causal links between purpose and health. Our identification strategy exploits the relation between *changes* in life purpose, *changes* in health, and the three channels proposed to underlie a causal link between life purpose levels and future health and mortality— behavioral (e.g., higher purpose causes individuals to eat more fruit and vegetables or use more preventive health care),^19,20^ biological (e.g., higher purpose causes better lipid profiles or lower blood pressure),^19,21^ or stress-buffering (e.g., higher purpose causes individuals to better handle stress).^22,23^ Because the impact of these channels tends to be cumulative over respondent’s lives, these pathways should impact long-term morbidity and mortality outcomes at least as much as they impact near-term outcomes. For instance, work demonstrates that diet, lipids, blood pressure, and preventive health care are at least as strongly related to long-term mortality risks as near-term mortality risks.^24–27^ In short, if the purpose-mortality relation primarily arises from purpose causing longevity, then a change in purpose will cause a change in behavior, biology, or stress-buffering and these changes should impact health in the future at least as much as they impact health today.

Alternatively, if changes in health today cause a change in life purpose, then (1) changes in purpose will be more strongly related to contemporaneous changes in health than future changes in health, and (2) life purpose *levels* will reflect (i.e., be caused by), to some degree, cumulative lifetime health changes. As a result, purpose levels will be correlated with health levels and unless one is able to perfectly control for health levels, purpose will also be correlated with future longevity because purpose proxies for health levels.^28,29^

Consider a simple example—a person experiences an increase in purpose (either through intervention or some exogenous factor such as the birth of a grandchild) and therefore quits smoking. The tobacco cessation will likely impact their risk of having a stroke five years from today at least as much as their risk of having a stroke tomorrow.^30^ Thus, if purpose levels cause future morbidity and mortality risk then the relation between *changes* in purpose and *changes* in health (i.e., health shocks) should not substantially attenuate with time. In contrast, consider a person who suffers (and survives) a stroke today which causes both a decline in purpose today and greater mortality risk tomorrow. If the correlation between purpose levels and future health outcomes primarily arises from purpose levels proxying for current health levels, then changes in purpose will be more strongly related to changes in current health than changes in future health.

## Methods

### Study Design and Participants

The Health and Retirement Study (HRS) is a nationally representative biennial prospective cohort survey of individuals age 50 and older and their spouses.^31,32^ (HRS is sponsored by the National Institute on Aging, grant number U01AG009740, and is conducted by the University of Michigan.) Beginning in 2006, HRS randomly selected half the participants for an enhanced face-to-face interview and psychosocial leave-behind questionnaire. The sample rotates with each HRS wave such that those respondents selected in 2006 are again selected (along with any new cohorts) in 2010, 2014, and 2018. The leave-behind questionnaire includes the seven question Ryff and Keyes life purpose measure and the face-to-face interviews include physical measures of lung function, grip strength, balance, and for those 65 and over, walking speed.^33^ The biennial core HRS survey also asks respondents a series of questions regarding medical diagnoses since their previous interview including a *new* diagnosis of hypertension, diabetes, cancer, lung disease, heart problem, stroke, psychological problem, arthritis, dementia, and Alzheimer’s disease. Because self-rated health may be impacted by “psychological distress and well-being,”^7^ the 14 measures of changes in health are objective—either measured directly (e.g., grip strength) or diagnosed by a physician. The appendix (pp 1-5) provides details for all metrics. The study was exempted from University of Arizona IRB because it uses deidentified publicly available information.

The initial sample consists of 12 745 individuals over age 49 who were eligible for the leave behind questionnaire in 2006, 2010, or 2014. For the contemporaneous analysis (years 1-4), respondents must answer at least four life purpose questions in both the initial and subsequent interview and have covariate data at the beginning of year 1. Analogously, for the subsequent four-year period, respondents must answer at least four life purpose questions in both years 1 and 4, have covariate data at the beginning of year 5, and survive to the end of year 8. For the 9-12 year period, respondents must answer at least four life purpose question in both years 1 and 4, have covariate data at the beginning of year 9, and survive to the end of year 12. Appendix pp 6-11 provides data cleaning flowcharts.

### Procedures

Our approach focuses on changes in health and changes in purpose. For the 10 diagnoses-based health shocks, individuals with an initial diagnosis in the period under examination are compared to individuals without the condition at the end of the period under examination (e.g., individuals who had their first stroke in the 2010-2014 period versus individuals who had never suffered a stroke by their 2014 interview). For three of the four physical health measures—lung function, grip strength, and walking speed—each interview wave we sort individuals into age and sex groups and then, within each age-sex group, partition into three groups by the change in the metric. We then classify those individuals within the 1/3^rd^ of observations with the largest decrease in physical health (e.g., a large decline in lung function) as suffering a negative health shock and those individuals within the top 1/3^rd^ as the non-shock sample (recognizing that these individuals will often have an increase in the metric) and reaggregate over the sex-age groups. As detailed in the appendix (pp 1-2), the classification for the balance test is more complicated as it is a series of three tests that are conditional on success in the initial test. We limit the sample to those individuals who successfully completed the first and second balance test in the initial year. We classify those that successfully completed the first and second balance test in the subsequent interview as the non-shock sample and those that fail or did not attempt (because they felt unsafe or were medically disqualified) the first test in the subsequent interview as the negative health shock sample.

Our analyses include the full set of covariates (except an indicator variable for the presence of a chronic disease because the initial diagnosis of these diseases defines some of our health shocks) used in a recent high-quality study linking purpose to longevity.^1^ Specifically, this set of covariates (see appendix pp 4-5) includes age, sex, education levels, race, marital status, smoking status, physical activity, alcohol frequency, and a functional health score. Because changes in purpose are negatively related to initial purpose levels (e.g., an individual with life purpose of 6 at the beginning of the period can only have a negative change in life purpose), all analyses include initial life purpose levels as a covariate.

### Statistical analysis

Our initial tests regress changes in life purpose over a four-year window on the control variables and an indicator for a health shock over the same four-year window. To interpret these results as health shocks causing life purpose requires that at least some of the change in purpose occurs because of the health shock. It is possible, however, that a change in purpose causes a near-term change in the likelihood of a health shock, e.g., a decline in purpose in year 1 causes an increased likelihood of a stroke in years 2-4. If causation primarily runs from purpose to future health shocks, however, then the associated changes in behavior, biology, or stress buffering should cause a decline in future health shocks at least as much as they cause a decline in “contemporaneous” health shocks. Thus, we next focus on logistic regressions that use the change in purpose in years 1-4 to explain variation in health shocks in years 1-4, 5-8, and 9-12. Because respondents can appear multiple times, standard errors are clustered at the respondent level. All analyses were conducted from December 15, 2021 to February 15, 2022, using SAS version 9·4. A two-tailed *p* value<0·05 was considered statistically significant.

## Results

Pooling over the three periods (2006-2010, 2010-2014, 2014-2018), the first two columns in Table 1 reports the number of individuals suffering the negative health shock and the number of individuals not suffering the shock by the end of the period. For example, the fifth row shows that 1 099 individuals were diagnosed with hypertension sometime over the 2006-2010, 2010-2014, or 2014-2018 period while 5 788 individuals were included in at least one of three periods and had never been diagnosed with hypertension. Analogously, the middle two columns report that, of those individuals with changes in purpose data in years 1-4 (and other covariates), 512 individuals were diagnosed with hypertension in years 5-8 and 2 932 had never been diagnosed with hypertension by the end of year 8. The final two columns report that, of those respondents with changes in purpose data in years 1-4, 180 were diagnosed with hypertension in years 9-12 and 955 had never been diagnosed with hypertension by the end of year 12. The appendix (pp 12-15) provides sample details.

**Table 1:**
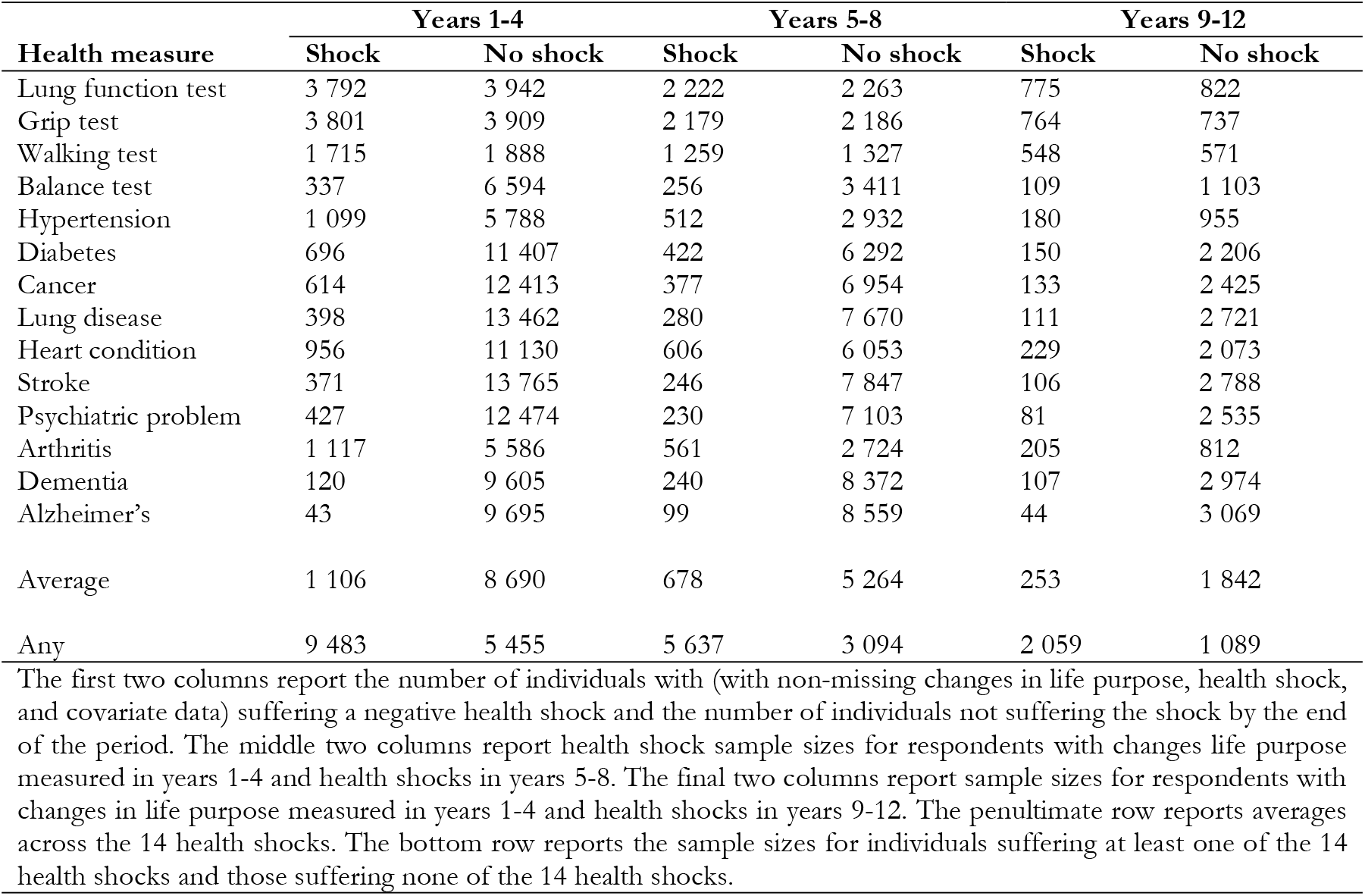
Health shock sample sizes.

Table 2 presents panel regressions of standardized (rescaled to unit variance and zero mean) changes in purpose in years 1-4 on the health shock indicator variable over the same four-year period and beginning of period covariates. The coefficients reflect the standard deviation change in purpose associated with that health shock controlling for respondent characteristics. The first row reveals, for instance, that relative to the 1/3^rd^ of individuals who suffer the smallest decline in lung function, the 1/3^rd^ of individuals who suffer the largest decline lung function contemporaneously suffer an 8·1% standard deviation decline in life purpose (*p*<0·001). Analogously, the fifth row reveals that relative to individuals who had not been diagnosed with hypertension by the end of the four-year period, individuals who experience their initial hypertension diagnosis suffer a 6·6% standard deviation decline in life purpose (*p*<0·04). In total, the relation between health shocks and contemporaneous changes in life purpose is negative for 13 of the 14 cases, and the coefficient associated with the health shock is statistically significant in 12 of the 14 cases.

**Table 2:** Regression of changes in life purpose on contemporaneous health shocks.

|  | **Coefficient** | **p-value** | **N** |
| --- | --- | --- | --- |
| Lung function test | -0.081 | 0.001 | 7 734 |
| Grip test | -0.077 | 0.001 | 7 710 |
| Walking test | -0.078 | 0.009 | 3 603 |
| Balance test | -0.125 | 0.023 | 6 931 |
| Hypertension | -0.066 | 0.035 | 6 887 |
| Diabetes | 0.021 | 0.540 | 12 103 |
| Cancer | -0.041 | 0.241 | 13 027 |
| Lung disease | -0.195 | 0.001 | 13 860 |
| Heart condition | -0.087 | 0.006 | 12 086 |
| Stroke | -0.209 | 0.001 | 14 136 |
| Psychiatric problem | -0.270 | 0.001 | 12 901 |
| Arthritis | -0.062 | 0.036 | 6 703 |
| Dementia | -0.435 | 0.001 | 9 725 |
| Alzheimer's | -0.381 | 0.021 | 9 738 |
| Any of the above | -0.058 | 0.001 | 14 938 |
| Number of shocks | -0.041 | 0.001 | 14 938 |

This table reports coefficients associated with a negative health shock in years 1-4 from regressions of standardized (rescaled to unit variance) change in life purpose in year 1-4 on negative health shocks over the same period and additional covariates including initial life purpose level, age, sex, education, marital status, smoking status, frequency of vigorous physical activity, days/week consuming alcohol, body mass index, and functional status.

The penultimate row reports the analysis when the health shock variable identifies individuals who experienced any of the 14 health shocks relative to individuals who did not experience any of the 14 health shocks over the four-year period. The bottom row reports an analogous regression but replaces the health shock metric with the number of health shocks a subject experienced over the four-year period. Both regressions continue to support the hypothesis that a health shock causes a decline in purpose, e.g., the bottom row suggests that each additional health shock is associated with a 4·1% standard deviation decline in purpose.

The results in Table 2 reveal that either negative health shocks cause substantial changes in life purpose or a change in purpose causes a near-immediate change in the likelihood of experiencing a health shock. Table 3 helps differentiate these explanations as it reports logistic regression results for (1) a health shock in the same period as the change in life purpose (e.g., both measured over 2006-2010; first three columns), (2) a health shock in period after the change in life purpose (e.g., health shock in 2010-2014 on the change in life purpose in 2006-2010; middle three columns), and (3) a health shock two periods after the change in life purpose (health shock in 2014-2018 on the change in life purpose in 2006-2010; final three columns). All analyses include the full set of control variables. Because the first three health shocks arise from the top or bottom third of changes in the measure, we also include the beginning of period health level as a covariate in these cases, e.g., the regression used to estimate the odds ratio for a decline in lung function over years 1-4 includes the lung function measure at the beginning of year 1. The remaining health shock samples are limited to individuals who are not diagnosed with the condition prior to the health shock period, e.g., the sample for the final three columns in the “cancer” row is limited to individuals who had never had a cancer diagnosis by the beginning of year 9.

**Table 3:** Changes in life purpose (years 1-4) and contemporaneous (years 1-4) and subsequent (years 5-8 and years 9-12) health shocks.

|  | Health shock in years 1-4 |  |  | Health shock in years 5-8 |  |  | Health shock in years 9-12 |  |  |
| --- | --- | --- | --- | --- | --- | --- | --- | --- | --- |
|  | OR | (95% CI) | N | OR | (95% CI) | N | OR | (95% CI) | N |
| Lung function test<sup>a</sup> | 0.866 | (0.815-0.920) | 7 734 | 0.985 | (0.902-1.076) | 4 485 | 1.013 | (0.874-1.174) | 1 597 |
| Grip test<sup>a</sup> | 0.866 | (0.816-0.920) | 7 710 | 0.921 | (0.838-1.011) | 4 365 | 1.165 | (0.988-1.374) | 1 501 |
| Walking test<sup>a</sup> | 0.887 | (0.81-0.970) | 3 603 | 0.985 | (0.873-1.110) | 2 586 | 0.897 | (0.749-1.074) | 1 119 |
| Balance test<sup>b</sup> | 0.847 | (0.723-0.991) | 6 931 | 0.810 | (0.662-0.991) | 3 667 | 0.889 | (0.671-1.177) | 1 212 |
| Hypertension<sup>c</sup> | 0.906 | (0.827-0.992) | 6 887 | 1.021 | (0.887-1.176) | 3 444 | 1.007 | (0.803-1.264) | 1 135 |
| Diabetes<sup>c</sup> | 1.033 | (0.935-1.142) | 12 103 | 0.912 | (0.795-1.047) | 6 714 | 0.911 | (0.707-1.174) | 2 356 |
| Cancer<sup>c</sup> | 0.939 | (0.845-1.044) | 13 027 | 0.964 | (0.832-1.116) | 7 331 | 0.924 | (0.701-1.219) | 2 558 |
| Lung disease<sup>c</sup> | 0.763 | (0.671-0.869) | 13 860 | 0.798 | (0.680-0.936) | 7 950 | 0.916 | (0.698-1.201) | 2 832 |
| Heart condition<sup>c</sup> | 0.882 | (0.807-0.964) | 12 086 | 1.004 | (0.897-1.124) | 6 659 | 0.970 | (0.797-1.181) | 2 302 |
| Stroke<sup>c</sup> | 0.744 | (0.644-0.859) | 14 136 | 0.822 | (0.687-0.983) | 8 093 | 0.864 | (0.664-1.126) | 2 894 |
| Psychiatric problem<sup>c</sup> | 0.672 | (0.589-0.766) | 12 901 | 0.946 | (0.786-1.139) | 7 333 | 1.041 | (0.772-1.405) | 2 616 |
| Arthritis<sup>c</sup> | 0.909 | (0.834-0.991) | 6 703 | 1.033 | (0.906-1.179) | 3 285 | 0.847 | (0.671-1.071) | 1 017 |
| Dementia<sup>c</sup> | 0.558 | (0.441-0.705) | 9 725 | 0.765 | (0.633-0.925) | 8 612 | 0.938 | (0.719-1.223) | 3 081 |
| Alzheimer's<sup>c</sup> | 0.609 | (0.405-0.915) | 9 738 | 0.881 | (0.657-1.182) | 8 658 | 0.953 | (0.630-1.442) | 3 113 |
| Any | 0.913 | (0.873-0.956) | 14 938 | 1.007 | (0.947-1.071) | 8 731 | 0.996 | (0.895-1.108) | 3 148 |

The results in the first three columns are consistent with the regression analysis in revealing a strong relation between health shocks and contemporaneous changes in purpose—as 13 of the 14 shocks have odds ratios less than one, and 12 of the 14 are statistically significant. For instance, a one unit increase in the Ryff and Keyes purpose metric decreases the likelihood of being in the 1/3^rd^ of individuals who suffer the largest decline in lung function (relative to the 1/3^rd^ of individuals who experience the smallest decline in lung function) by 13% (OR=0·866, 95% CI 0·815-0·920). The bottom row in the first three columns reports the logistic regression where the dependent variable is one if the person experiences any of the 14 health shocks over the four-year period and demonstrates a strong contemporaneous negative relation between health shocks and change in purpose.

The middle three columns report the logistic regression of a health shock in the subsequent four-year period (years 5-8) on the change in purpose in the initial four-year period (years 1-4), the initial purpose level (at the beginning of year 1), and the other covariates measured at the beginning of year 5. Thus, for example, this test examines if an increase in purpose in 2006-2010 period causes behavioral, biological, or stress-buffering changes that in turn can help explain who is more likely to suffer a decline in lung function in the 2010-2014 period. The results reveal a much weaker relation between changes in purpose and health shocks over the subsequent four years as three of the 14 variables have odds ratios greater than 1 and only four of the odds ratios that are less than 1 are statistically meaningful. The bottom row reports the results of an analogous logistic regression where the dependent variable is an indicator for at least one of the 14 health shocks in years 5-8 on the same variables. In contrast to the contemporaneous results, there is no evidence a decrease in purpose is associated with an increased likelihood of suffering a health shock (OR=1·007, 95% CI 0·947-1·071) in the subsequent four-year period.

The final three columns report results for health shocks in the subsequent 9-12 years on covariates at the beginning of year 9, purpose at the beginning of year 1, and changes in purpose in years 1-4. There is no evidence that changes in purpose lead to reduced health shock risk as none of the odds ratios differ meaningfully from one. Similarly, the bottom row reveals no evidence that a person with a decrease in life purpose in years 1-4 is less likely to suffer at least one of the 14 health shocks in years 9-12.

## Discussion

There is strong evidence that negative health shocks cause a decline in purpose (Table 2 and first three columns of Table 3). Given a negative health shock, by definition, causes increased future morbidity and mortality risk, the results suggest that reverse causation plays a meaningful role in driving the previously documented relation between purpose levels and subsequent mortality. Importantly, the contemporaneous relation between changes in health and changes in purpose mean that purpose levels will reflect, to some degree, cumulative previous changes in health and therefore, even absent any causal relation, will proxy for current health levels and be associated with future mortality unless one perfectly controls for current health levels.

In contrast to the strong contemporaneous relation, results in the final six columns of Table 3 reveal little evidence that a change in purpose leads to changes in behavior, biology, or stress-buffering that, in turn, systematically influence the likelihood of suffering a future health shock. Our results may also be interpreted as a test of how purpose interventions may influence future health—whether the change in life purpose is organic (our sample) or through an intervention—there is little systematic evidence to suggest a meaningfully impact on future health.

Note also that even with our proposition that a change in health impacts life purpose (i.e., our contemporaneous finding), reverse causation may still cause a negative relation between changes in life purpose and *future* health shocks. For instance, if a decline in health precedes a lung disease diagnosis, one should expect a decline in purpose will also precede (but not cause) a lung disease diagnosis. In short, any link between changes in purpose and subsequent health outcomes can still suffer from reverse causation if health shocks cause changes in purpose.

In sum, our evidence suggests that the relation between purpose levels and future morbidity or mortality primarily arises because purpose levels proxy for current health. Thus, an alternative approach is to better control for current health and exclude individuals who are already ill (and are therefore more likely to die soon) from the analysis. In a companion study,^34^ we find that this alternative test yields the same conclusion—strong evidence that reverse causation plays the primary role in driving the previously well-documented purpose-mortality relation.

Finally, our results do not imply that purpose is unimportant or has zero causal relation with future health. Rather our evidence suggests that tests linking life purpose levels to future mortality and morbidity appear to primarily result from reverse causation.

### Limitations

Our sample was limited to individuals 50 and older. The relations between changes in purpose and contemporaneous and future health shocks may differ for a younger population. Moreover, given 10 of the 14 health shocks are based on respondents’ recollection of whether they had received a diagnosis since their prior interview, there is some measurement error. In addition, because we pool data (to ensure meaningful sample sizes) over four survey waves and 12 years (2006, 2010, 2014, and 2018), we equally weight observations. Although the relation between changes in life purpose and health shocks is primarily contemporaneous, to the extent that HRS oversamples some populations (Blacks, Hispanics, and Floridians) our estimates do not necessarily reflect population estimates.

It is also possible that future health benefits associated with higher purpose are not captured by the 14 objectively measured health metrics we examine. Moreover, many factors (e.g., genetic variation) give rise to health shocks that have nothing to do with life purpose which could imply that our tests may be underpowered to identify how changes in purpose cause changes in future health.

### Conclusions

There is strong evidence that health shocks cause a decline in purpose and, as a result, purpose levels will reflect cumulative health shocks. In contrast, there is little evidence that changes in purpose cause changes in behaviors, biology, or stress-buffering that in turn impact the likelihood of suffering from a future health shock. The evidence suggests that reverse causation—a health shock causes both greater mortality risk and lower purpose—plays a substantial role in the relation between life purpose and mortality risk.

## Data Availability

All data were derived from publicly available Health and Retirement Study and detailed in the supplemental appendix.

https://hrs.isr.umich.edu/

## Contributors

This study was designed by RS and HT. Both authors accessed and verified the data. Both authors were involved with data interpretation and contributed to the methods development. RS did the data and statistical analyses. Both authors contributed to drafting the paper and revised the manuscript for important intellectual content. Both authors approved the decision to submit the final version of the manuscript.

## Data sharing

All data were derived from publicly available sources as detailed in the supplemental appendix.

## Acknowledgements

We are grateful for the support of the University of Arizona and Colorado State University.

