## Supplemental appendix for "Health shocks and changes in life purpose: Understanding the link between purpose and longevity"

### S1. Health shocks

*Background for Physical Measurements*: Beginning in 2006, HRS began an assessment of physical performance for respondents selected for the face-to-face interview (as detailed in the study, a rotating sample of approximately half of HRS respondents are selected for enhanced face-to-face interviews every four years, e.g., those selected in 2006 are also selected in 2010, 2014, and 2018). Respondents in nursing homes, those who had a proxy respondent, and those who were only able to do a telephone interview were not given the physical tests. Respondents were asked to read and waive a consent form and, prior to each test, were asked if they understood the directions and felt safe. Those who did not understand or did not feel safe were not given the test. The process included nine physical measurements: blood pressure, pulse, lung function, hand grip strength, balance tests, timed walk test (for those age 65 and older), height, weight, and waist circumference. Given blood pressure is often controlled by medications, and pulse, or weight loss/gain may indicate either health improvement or health deterioration (and height is effectively constant), we limit the physical measures to the lung function, grip, balance, and timed walk tests.

*Lung Function Test*: Interviewers used a Mini-Wright Peak Flow Meter with a disposable mouthpiece to measure peak expiratory flow (the amount and rate that air can be pushed out of the lungs after a full inhalation) in liters per minute. Respondents were given three attempts (with at least 30 seconds between attempts). We use the variables RwPUFF (the maximum value from the three tests) for 2006 (wave, w=8), 2010 (w=10), 2014 (w=12) and 2018 (w=14) from the RAND 1992-2018 longitudinal file. For each wave, we sort respondents who have lung tests in adjacent waves, by sex and age group (50-54, 55-59, 60-64, 65-69, 70-74, 75-79, 80+) and then partition those within each sex/age group into three groups by the change in peak expiratory flow over the period. For instance, we sort respondents in 2006 into sex and (2006) age groups and, within each group, partition into terciles by the change in peak expiratory flow between 2006 and 2010. We repeat the process for the two subsequent periods (e.g., sort into 2010 age and sex groups and measure the change in lung function over 2010-2014). Those respondents in the bottom tercile each period are classified as having a negative health shock while those in the top tercile are classified as the non-shock group.

*Grip Test*: Using a "Smedley spring-type" hand dynamometer, respondents are asked to squeeze the dynamometer as hard as they can. Respondents are given four attempts—two attempts for their dominant hand and two attempts for their non-dominant hand. We use the variables RwGRP (the maximum value from the four grip tests) for 2006 (w=8), 2010 (w=10), 2014 (w=12) and 2018 (w=14) from the RAND 1992-2018 longitudinal file. We follow a process identical to that for lung function (see above for detail) to form age and sex stratified terciles of respondents for negative health shock and non-shock samples based on changes in grip strength.

*Walking Test*: Respondents age 65 and older are asked to walk at "normal pace" a 98.5 inch (2.5 meters) course (marked with masking tape). The interviewer used a stopwatch to time the walk and complete the test twice (timing the walk from point A to B and then timing it a second time from point B to point A). We use the variables RwTIMWLK (the minimum value in seconds to two decimal places for the two attempts) for 2006 (w=8), 2010 (w=10), 2014 (w=12) and 2018 (w=14) from the RAND 1992-2018 longitudinal file. We follow a process identical to that for lung function (see lung function test for detail) to form age and sex stratified terciles of respondents for negative health shock and non-shock samples based on changes in walking speed.

*Balance Test*: The balance tests consist of three tests—a semi-tandem stand, a side-by-side stand, and a full-tandem stand (see https://www.youtube.com/watch?v=_uG63lguulI for details of these tests). Respondents are first asked to hold a semi-tandem stand for 10 seconds. Respondents who failed to hold the semi-tandem stand for at least 10 seconds are asked to complete the side-by-side tandem stand for 10 seconds. If the respondent was able to hold the semi-tandem stand for 10 seconds, the respondent is asked to hold a full-tandem stand for 60 seconds (if age <70) or 30 seconds (if age 70 or older). We begin by limiting the sample to those respondents who, at the beginning of each period under evaluation, were able to hold the semi-tandem stand for at least 10 seconds and the full-tandem stand for 60 seconds (if age<70) or 30 seconds (if age 70 or older). That is, respondents who began the period passing both balance tests (because these respondents passed the semi-tandem test, they were not asked to perform the side-by-side tandem stand). We classify those respondents who pass both the semi-tandem and full-tandem tests in the subsequent interview as the no-shock sample. We classify respondents who fail the semi-tandem test in the subsequent interview (and therefore did not qualify to take the full tandem test in the subsequent interview) as well as respondents who were reported to attempted and failed the test (coded .W), felt unsafe and did not attempt the test (coded .R), or were medically disqualified from taking the test (coded .X) as the negative shock sample. We use the variables RwBALSEMI and RwBALFULT for 2006 (w=8), 2010 (w=10), 2014 (w=12) and 2018 (w=14) from the RAND 1992-2018 longitudinal file.

*Hypertension Diagnosis*: At each core interview wave (i.e., every two years), respondents are asked about a hypertension diagnosis. If it is the respondent's first interview, they are asked, "Has a doctor ever told you that you have high blood pressure or hypertension?" If it is not the respondent's first interview and the respondent previously reported they had hypertension, the respondent is told, "Our records from your last interview [in [previous interview wave month] [previous interview wave year]] show that you have had high blood pressure or hypertension" but interviewers record if the respondent disputes this information. If a respondent was previously interviewed and did not report hypertension, the respondent is asked, "Since we last talked to you [in [previous interview wave month] [previous interview wave year]], has a doctor told you that you have high blood pressure or hypertension?" Similarly, respondents are allowed to dispute the previous record. We use the variables RwHIBPE for 2006 (w=8), 2010 (w=10), 2014 (w=12) and 2018 (w=14) from the RAND 1992-2018 longitudinal file. When the respondent does not dispute the record, the RwHIBPE variable is set to yes (1) if the respondent reported a hypertension diagnosis in the current or any previous wave and no (0) if the respondent reported no hypertension diagnosis in the current or previous waves. The RAND variable sets the value no for all previous waves if a respondent disputes a previous record of hypertension. We define those respondents who report no hypertension diagnosis at both the beginning (e.g., 2006) and end (e.g., 2010) of the four-year period as the non-shock sample. We define respondents who report no hypertension diagnosis at the beginning of the period (e.g., 2006) but a hypertension diagnosis at the end of the period (e.g., 2020) as the negative health shock sample.

*Diabetes Diagnosis*: Analogous to hypertension diagnosis. Respondents are asked, "…has a doctor told you that you have diabetes or high blood sugar?" We use the variables RwDIABE for 2006 (w=8), 2010 (w=10), 2014 (w=12) and 2018 (w=14) from the RAND 1992-2018 longitudinal file.

*Cancer Diagnosis*: Analogous to hypertension diagnosis. Respondents are asked, "…has a doctor told you that you have cancer or a malignant tumor, excluding minor skin cancer?" We use the variables RwCANCRE for 2006 (w=8), 2010 (w=10), 2014 (w=12) and 2018 (w=14) from the RAND 1992-2018 longitudinal file.

*Lung Disease Diagnosis*: Analogous to hypertension diagnosis. Respondents are asked, "…has a doctor told you that you have chronic lung disease such as chronic bronchitis or emphysema?" We use the variables RwLUNGE for 2006 (w=8), 2010 (w=10), 2014 (w=12) and 2018 (w=14) from the RAND 1992-2018 longitudinal file.

*Heart Condition Diagnosis*: Analogous to hypertension diagnosis. Respondents are asked, "…has a doctor told you that you have had a heart attack, (have) coronary heart disease, angina, congestive heart failure, or other heart problems?" We use the variables RwHEARTE for 2006 (w=8), 2010 (w=10), 2014 (w=12) and 2018 (w=14) from the RAND 1992-2018 longitudinal file.

*Stroke Diagnosis*: Analogous to hypertension diagnosis. Respondents are asked, "…has a doctor told you that you have had a stroke?" We use the variables RwSTROKE for 2006 (w=8), 2010 (w=10), 2014 (w=12) and 2018 (w=14) from the RAND 1992-2018 longitudinal file.

*Psychiatric Problem Diagnosis*: Analogous to hypertension diagnosis. Respondents are asked, "…have you had or has a doctor told you that you have any emotional, nervous, or psychiatric problems?" We use the variables RwPSYCHE for 2006 (w=8), 2010 (w=10), 2014 (w=12) and 2018 (w=14) from the RAND 1992-2018 longitudinal file.

*Arthritis Diagnosis*: Analogous to hypertension diagnosis. Respondents are asked, "…have you had or has a doctor told you that you have Arthritis or rheumatism?" We use the variables RwARTHRE for 2006 (w=8), 2010 (w=10), 2014 (w=12) and 2018 (w=14) from the RAND 1992-2018 longitudinal file.

*Dementia Diagnosis*: Analogous to hypertension diagnosis. Respondents are asked, "…has a doctor told you that you have Dementia, senility or any other serious memory impairment?" We use the variables RwDEMENE for 2010 (w=10), 2014 (w=12) and 2018 (w=14) from the RAND 1992-2018 longitudinal file (this question is first asked in the 2010 wave).

*Alzheimer's Disease Diagnosis*: Analogous to hypertension diagnosis. Respondents are asked, "…has a doctor told you that you have Alzheimer's Disease?" We use the variables RwALZHE for 2010 (w=10), 2014 (w=12) and 2018 (w=14) from the RAND 1992-2018 longitudinal file (this question is first asked in the 2010 wave).

### S2. *Supplementary Table 1:* Variable creation of life purpose and control variables

| Variable | Variable ID/Construction | Source |
|---|---|---|
| Leave behind questionnaire eligibility | KLBELIG (2006) MLBELIG (2010), OLBELIG (2014), QLBELIG (2018) | 2006, 2010, 2014 from RAND fat files; 2018 from raw HRS data (h18lg_r) |
| Life purpose score | For individuals who answer at least four of the seven questions, e.g., for 2006:<br>LP=sum(KLB035A,-1*KLB035B,KLB035C,-1*KLB035D, -1*KLB035E,-1*KLB035F,KLB035G) | RAND 2006, 2010, 2014, and 2018 fat files |
| Age (years) | r8agey_b, r10agey_b, r12agey_b – Respondent current age calculation in 2006, 2010, or 2014; form age groups 50-54, 55-59, 60-64, 65-69, 70-74, 75-79, 80+ | RAND longitudinal file |
| Sex | RAGENDER | RAND longitudinal file |
| Marital status | RwMSTAT<br>Married: RwMSTAT=1, 2, or 3<br>Divorced: RwMSTAT=4, 5, or 6<br>Widowed: RwMSTAT=7<br>Never married: RwMSTAT=8 | RAND longitudinal file |
| Race/ethnicity | RACACEM and RAHISPAN<br>White: RACACEM=1 and RAHISPAN=0<br>Black: RACEM=2<br>Hispanic White: RACACEM=1 RAHISPN=1<br>Other: RARACEM=3 | RAND longitudinal file |
| Education level | RAEDUC and RAEDEGRM<br>Less than high school: RAEDUC=1<br>High school graduate: RAEDUC=2 or RAEDUC=3<br>Some college: RAEDUC=4<br>College: RAEDUC=5 and RAEDEGRM NE 6 or 7<br>Graduate degree: RAEDEGRM=6 or 7 | RAND longitudinal file |
| Smoking status | RwSMOKEN and RwSMOKEV<br>Current smoker: RwSMOKEN=1<br>Former smoker: RwSMOKEN NE 1 and RwSMOKEV=1<br>Never smoker: RwSMOKEV=0 | RAND longitudinal file |
| Alcohol consumption | RwDRINKD<br>0: RwDRINKD=0<br>1-2: RwDRINKD=1 or RwDRINKD=2<br>3-4: RwDRINKD=3 or RwDRINKD=4<br>5-6: RwDRINKD=5 or RwDRINKD=6<br>7: RwDRINKD=7 | RAND longitudinal file |
| BMI | RwBMI<br>Low BMI: RwBMI<18.5<br>Normal BMI: 18.5 <=RwBMI<25<br>Overweight BMI: 25<=RwBMI<30<br>Obese BMI: RwBMI>30 | RAND longitudinal file |

| Variable | Variable ID/Construction | Source |
| --- | --- | --- |
| Vigorous physical exercise | RwVGACTX<br>Daily: RwVGACTX=1<br>>1/week: RwVGACTX=2<br>1/week: RwVGACTX=3<br>1-3/month: RwVGACTX=4<br>Hardly ever or never: RwVGACTX=5 | RAND longitudinal file |
| Functional score | RwADLA | RAND longitudinal file |

### S3. *Supplementary Figure 1:* 2006-2010 Contemporaneous cleaning flowchart

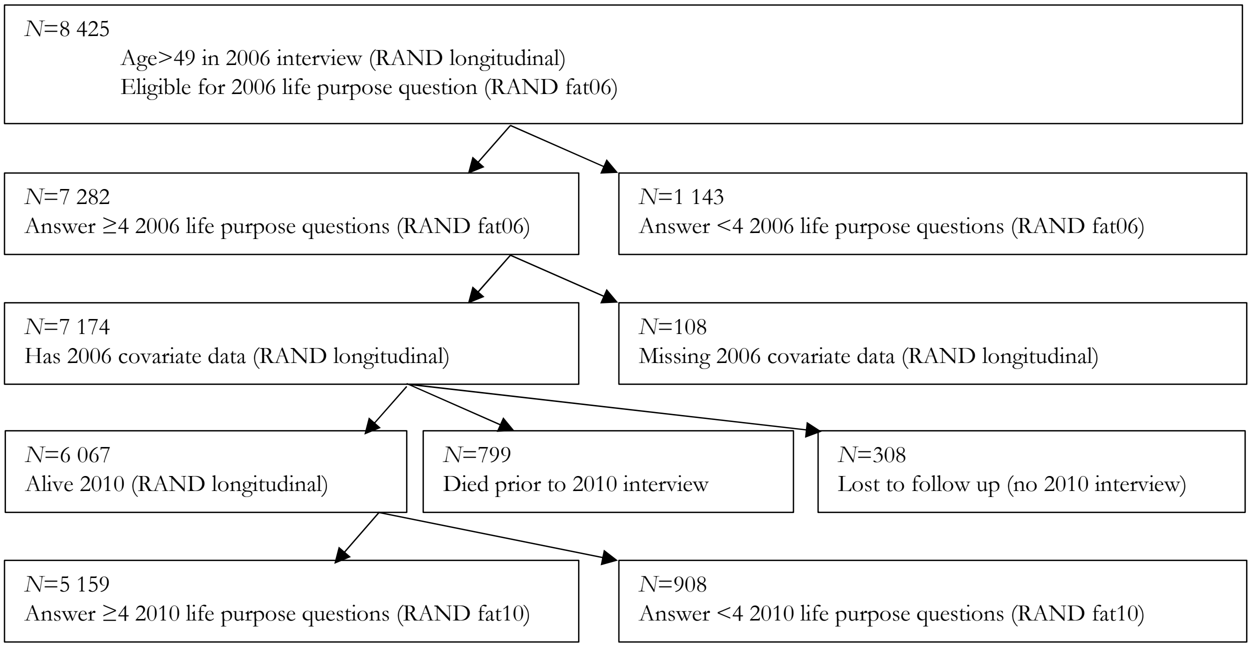

### S4. ***Supplementary Figure 2:*** 2010-2014 Contemporaneous cleaning flowchart

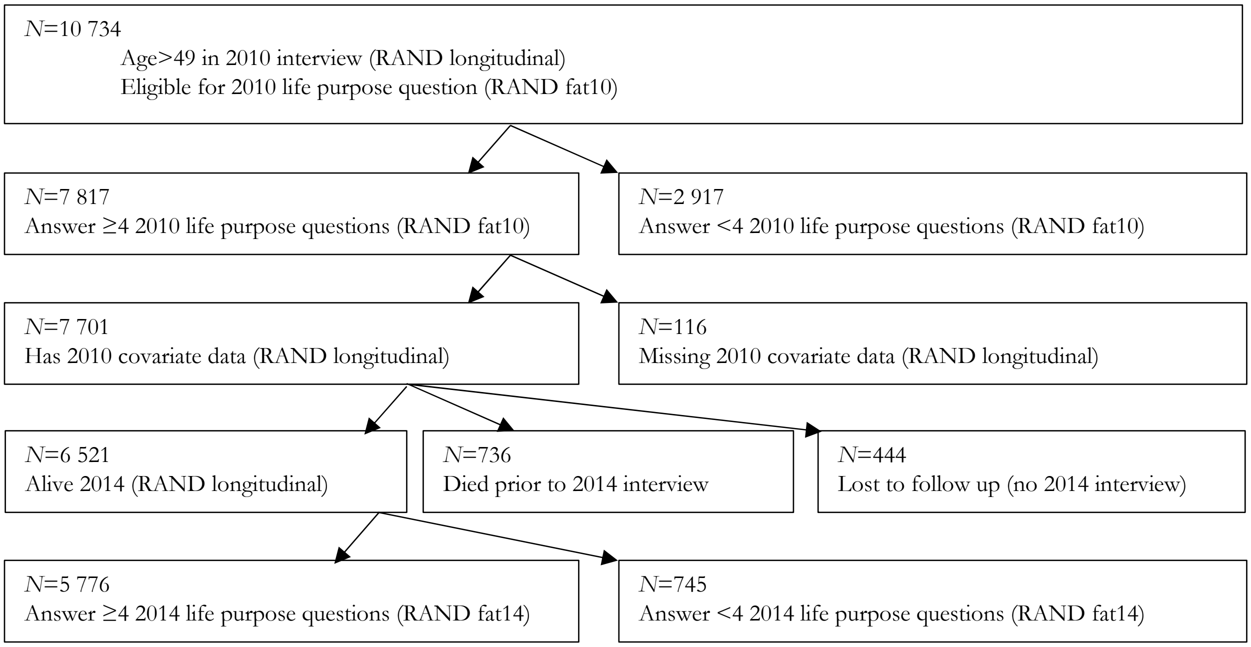

### S5. *Supplementary Figure 3:* 2014-2018 Contemporaneous cleaning flowchart

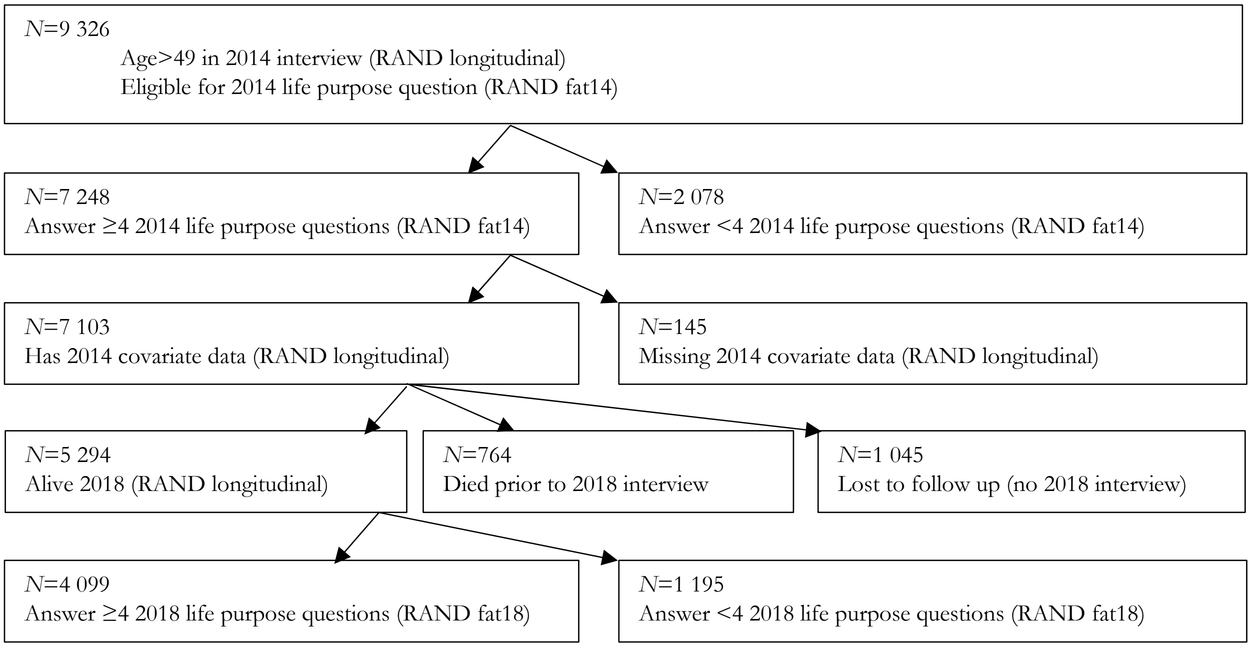

**S6. *Supplementary Figure 4:* 2010-2014 Health shock and 2006-2010 change in life purpose flowchart**

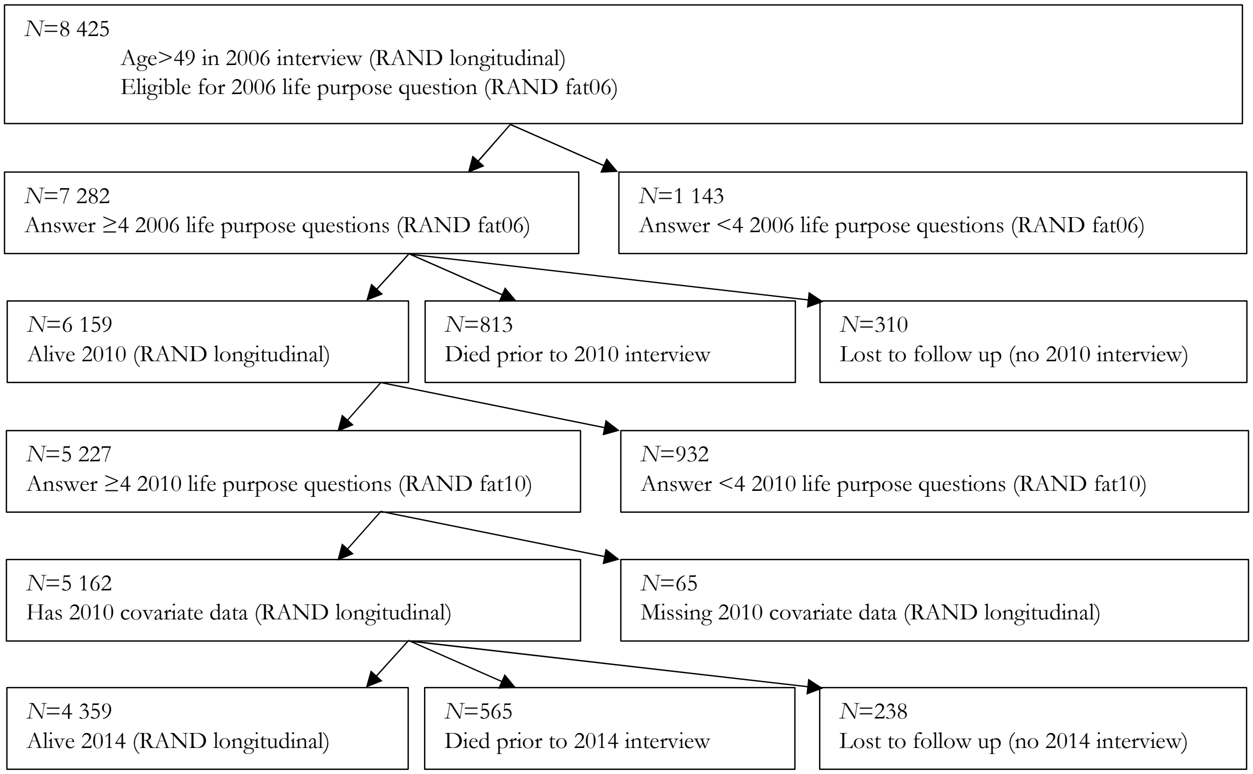

### S7. *Supplementary Figure 5:* 2014-2018 Health shock and 2010-2014 change in life purpose flowchart

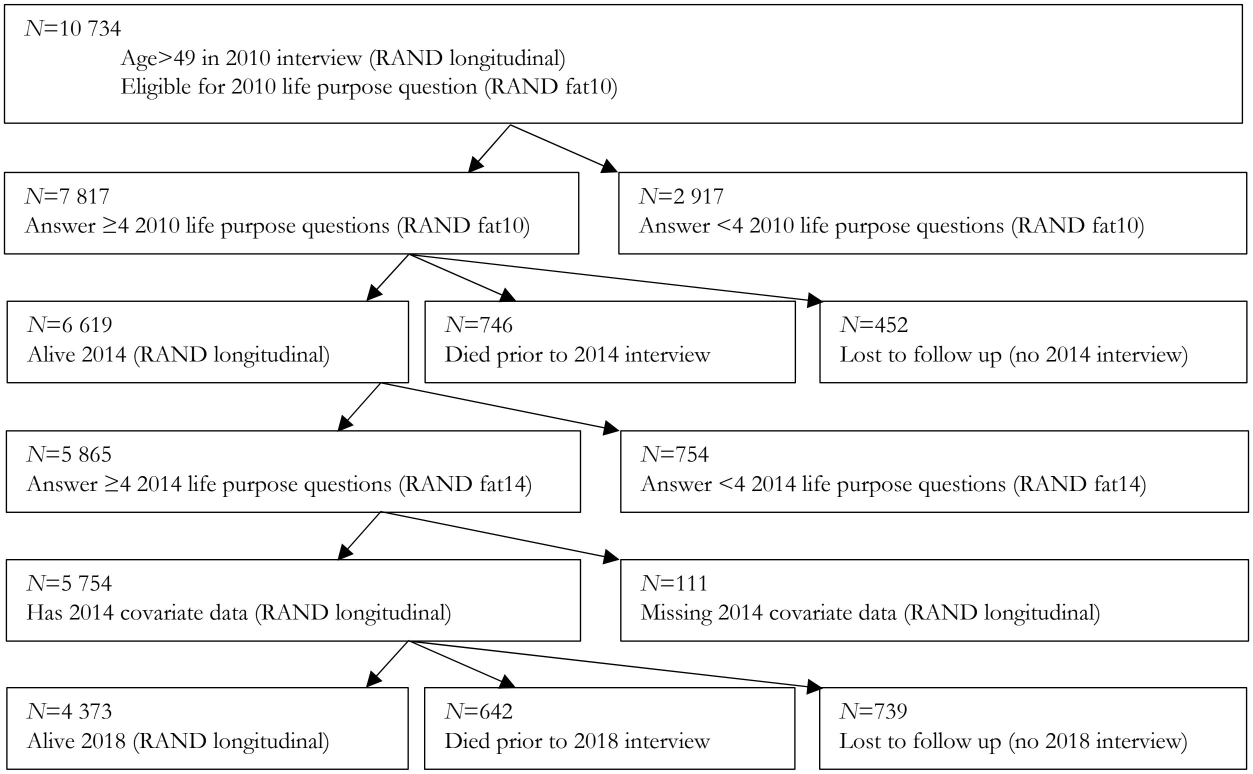

**S8. *Supplementary Figure 6:* 2014-2018 Health shock and 2006-2010 change in life purpose flowchart**

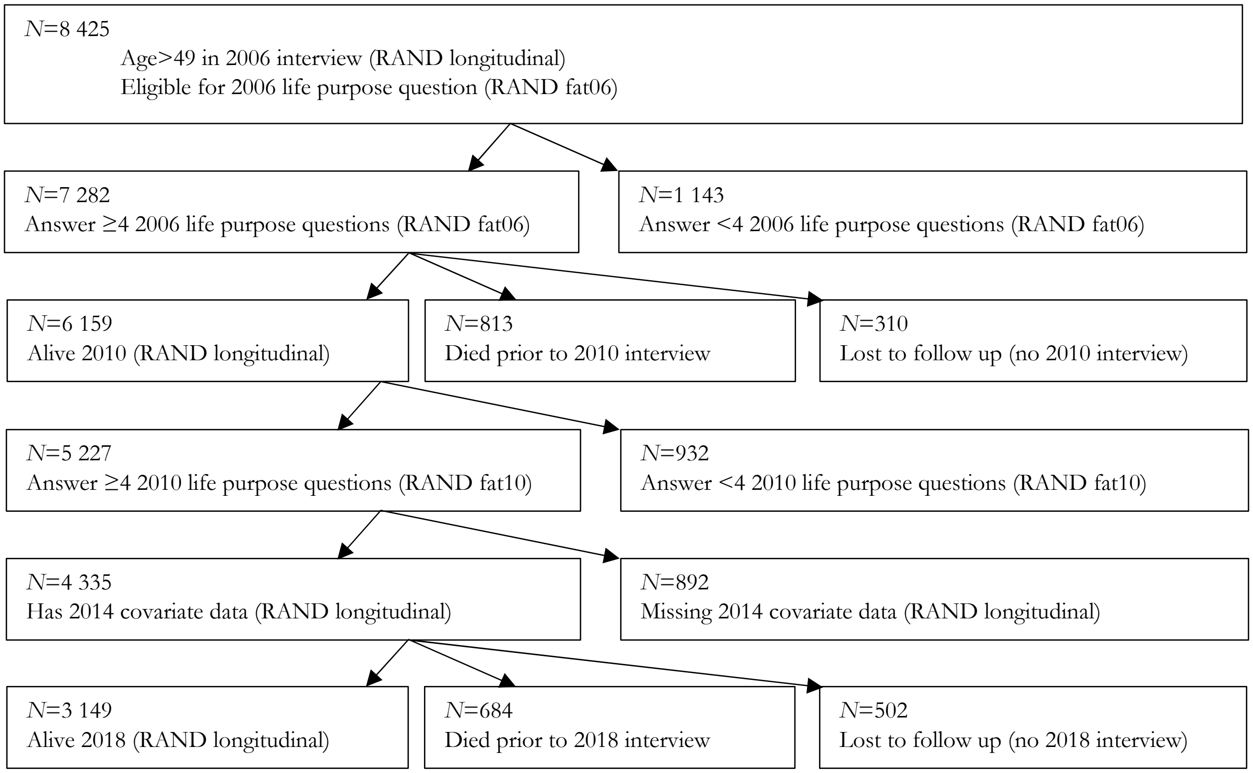

### S9. ***Supplementary Table 2:*** Sample construction details

Because we focus on health shocks that occur over a period and changes in life purpose, our sample consists of three periods (2006-2010, 2010-2014, and 2014-2018). The first three columns in the first row of Supplementary Table 2 report the raw (prior to conditioning on health conditions) sample size for the contemporaneous tests (i.e., examination of health shocks and changes in life purpose over the same four-year period). Thus, the values in the first three cells are identical to the values in the bottom cells of cleaning flowcharts S3, S4, and S5, respectively. The penultimate column reports the total sample size (i.e., the sum of the previous three cells) and the final column reports the number of unique individuals in the sample. The second row reports analogous figures for the sample that considers changes in life purpose in years 1-4 and health shocks in years 5-8 (i.e., are identical to the bottom left cells in flowcharts S6 and S7, respectively). Analogously, the third row reports figures for the sample that considers changes in life purpose in years 1-4 and health shocks in years 9-12 (i.e., is identical to the bottom left cell in flowchart S8).

The remaining rows report the number of individuals, within each of the four-year periods, that have sufficient data to compute their change in health (e.g., had their grip strength measured at both the beginning and end of the four-year period) over the contemporaneous period (i.e., these values are subsets of the top row).

| | 2006-2010 | 2010-2014 | 2014-2018 | Total Observations | Unique Individuals |
|---|---|---|---|---|---|
| Contemporaneous | 5 159 | 5 776 | 4 099 | 15 034 | 7 598 |
| Years 5-8 | | 4 359 | 4 373 | 8 732 | 5 793 |
| Years 9-12 | | | 3 149 | 3 149 | 3 149 |
| Lung function test | 3 969 | 4 516 | 3 531 | 12 016 | 6 099 |
| Grip test | 3 908 | 4 369 | 3 404 | 11 681 | 5 995 |
| Walking test | 2 064 | 2 090 | 1 768 | 5 922 | 3 355 |
| Balance test | 4 108 | 4 608 | 3 615 | 12 331 | 6 196 |
| Hypertension | 5 148 | 5 764 | 4 090 | 15 002 | 7 585 |
| Diabetes | 5 152 | 5 769 | 4 091 | 15 012 | 7 588 |
| Cancer | 5 146 | 5 762 | 4 086 | 14 994 | 7 583 |
| Lung disease | 5 148 | 5 766 | 4 089 | 15 003 | 7 587 |
| Heart condition | 5 151 | 5 763 | 4 093 | 15 007 | 7 588 |
| Stroke | 5 153 | 5 766 | 4 095 | 15 014 | 7 590 |
| Psychiatric problem | 5 151 | 5 766 | 4 090 | 15 007 | 7 587 |
| Arthritis | 5 152 | 5 769 | 4 096 | 15 017 | 7 591 |
| Dementia | | 5 774 | 4 096 | 9 870 | 6 359 |
| Alzheimer's | | 5 756 | 4 094 | 9 850 | 6 343 |

The top three rows report sample sizes for estimates based on changes in life purpose in years 1-4 and health shock data in years 1-4 (i.e., contemporaneous), health shock data in years 5-8, and health shock data in years 9-12, respectively. The penultimate column is the sum of the first three columns and the final column reports the number of unique individuals in the sample. The bottom 14 rows report the available (contemporaneous) observations for the 14 change in health measures we consider for the three sample periods, followed by the total number of observations, and the unique number of individuals.

.

### S10. *Supplementary Table 3:* Contemporaneous health shocks sample by year

| | 2006-2010 | | 2010-2014 | | 2014-2018 | | Total | |
|---|---|---|---|---|---|---|---|---|
| | **Shock** | **No Shock** | **Shock** | **No Shock** | **Shock** | **No Shock** | **Shock** | **No Shock** |
| Lung function test | 1 283 | 1 347 | 1 461 | 1 465 | 1 048 | 1 130 | 3 792 | 3 942 |
| Grip test | 1 286 | 1 282 | 1 459 | 1 502 | 1 056 | 1 125 | 3 801 | 3 909 |
| Walking test | 666 | 703 | 661 | 716 | 388 | 469 | 1 715 | 1 888 |
| Balance test | 128 | 2 127 | 136 | 2 436 | 73 | 2 031 | 337 | 6 594 |
| Hypertension | 449 | 2 019 | 374 | 2 152 | 276 | 1 617 | 1 099 | 5 788 |
| Diabetes | 277 | 3 981 | 243 | 4 361 | 176 | 3 065 | 696 | 11 407 |
| Cancer | 233 | 4 234 | 240 | 4 764 | 141 | 3 415 | 614 | 12 413 |
| Lung disease | 162 | 4 614 | 159 | 5 173 | 77 | 3 675 | 398 | 13 462 |
| Heart condition | 366 | 3 785 | 369 | 4 226 | 221 | 3 119 | 956 | 11 130 |
| Stroke | 155 | 4 720 | 143 | 5 285 | 73 | 3 760 | 371 | 13 765 |
| Psychiatric problem | 149 | 4 385 | 163 | 4 777 | 115 | 3 312 | 427 | 12 474 |
| Arthritis | 406 | 1 849 | 410 | 2 140 | 301 | 1 597 | 1 117 | 5 586 |
| Dementia | | | 97 | 5 641 | 23 | 3 964 | 120 | 9 605 |
| Alzheimer's | | | 33 | 5 712 | 10 | 3 983 | 43 | 9 695 |

The final two columns match the sample sizes reported in Table 1 for years 1-4 and are the sum of the last two columns matches the sample sizes reported in third columns of Tables 2 and 3. The first six columns report the sample sizes by period. The sum of columns 1, 3, and 5 equals column 7 and the sum of columns 2, 4, and 6 equals column 8.

### S11. ***Supplementary Table 4:*** Health shock in years 5-8 by year (change in life purpose in years 1-4)

| | 2010-2014 | | 2014-2018 | | Total | |
|---|---|---|---|---|---|---|
| | **Shock** | **No shock** | **Shock** | **No shock** | **Shock** | **No shock** |
| Lung function test | 1 095 | 1 106 | 1 127 | 1 157 | 2 222 | 2 263 |
| Grip test | 1 089 | 1 109 | 1 090 | 1 077 | 2 179 | 2 186 |
| Walking test | 661 | 708 | 598 | 619 | 1 259 | 1 327 |
| Balance test | 114 | 1 653 | 142 | 1 758 | 256 | 3 411 |
| Hypertension | 266 | 1 481 | 246 | 1 451 | 512 | 2 932 |
| Diabetes | 176 | 3 219 | 246 | 3 073 | 422 | 6 292 |
| Cancer | 200 | 3 453 | 177 | 3 501 | 377 | 6 954 |
| Lung disease | 132 | 3 846 | 148 | 3 824 | 280 | 7 670 |
| Heart condition | 313 | 3 000 | 293 | 3 053 | 606 | 6 053 |
| Stroke | 131 | 3 898 | 115 | 3 949 | 246 | 7 847 |
| Psychiatric problem | 114 | 3 594 | 116 | 3 509 | 230 | 7 103 |
| Arthritis | 264 | 1 356 | 297 | 1 368 | 561 | 2 724 |
| Dementia | 134 | 4 174 | 106 | 4 198 | 240 | 8 372 |
| Alzheimer's | 58 | 4 262 | 41 | 4 297 | 99 | 8 559 |

The final two columns match the sample sizes reported in Table 1 for years 5-8 and the sum of the last two columns matches the sample size reported in sixth column of Table 3. The four first columns report the sample sizes by period. The sum of columns 1 and 3 equals column 5 and the sum of columns 2 and 4 equals column 6.

**S12.** ***Supplementary Table 5:*** **Health shocks in years 9-12 by year (change in life purpose in years 1-4)**

| | 2014-2018 | | Total | |
|---|---|---|---|---|
| | **Shock** | **No shock** | **Shock** | **No shock** |
| Lung function test | 775 | 822 | 775 | 822 |
| Grip test | 764 | 737 | 764 | 737 |
| Walking test | 548 | 571 | 548 | 571 |
| Balance test | 109 | 1 103 | 109 | 1 103 |
| Hypertension | 180 | 955 | 180 | 955 |
| Diabetes | 150 | 2 206 | 150 | 2 206 |
| Cancer | 133 | 2 425 | 133 | 2 425 |
| Lung disease | 111 | 2 721 | 111 | 2 721 |
| Heart condition | 229 | 2 073 | 229 | 2 073 |
| Stroke | 106 | 2 788 | 106 | 2 788 |
| Psychiatric problem | 81 | 2 535 | 81 | 2 535 |
| Arthritis | 205 | 812 | 205 | 812 |
| Dementia | 107 | 2 974 | 107 | 2 974 |
| Alzheimer's | 44 | 3 069 | 44 | 3 069 |

The final two columns match the sample sizes reported in Table 1 for years 9-12 and the sum of the last two columns matches the sample size reported in last column of Table 3. The two columns report the sample sizes by period. Because there is only one period—changes in life purpose in 2006-2010 and health shocks in 2014-2018, the first and third (and the second and fourth) columns are identical.

### S13. Related Studies

Our study examines the relation between changes in life purpose and both contemporaneous and subsequent changes in health. Surprisingly, given concerns regarding reverse causation, most previous work focuses on levels rather than changes. Nonetheless, some previous work (using various methodologies, datasets, and measures of life purpose) examine the relations between changes in health and previous life purpose levels,[1] health levels and both previous life purpose levels and health levels,[1] and changes in life purpose and lag health levels.[2] Work also examines outcomes (including health metrics) at the end of period t+2 on the change in life purpose between t and t+1 and the outcome level at time t.[2,3] In these cases, however, because the change in outcome is measured from t to t+2 while the change in life purpose is measured from t to t+1, one cannot differentiate contemporaneous versus subsequent changes in outcomes.

In sum, none of the approaches in the extant literature are equivalent to our "difference in difference" design that examines the relation between changes in life purpose and both contemporaneous and subsequent changes in health.

### S14. STROBE statement

Following Alijumiang et al,[4] and for the sake of brevity, the present study followed the Strengthening the Reporting of Observational Studies in Epidemiology (STROBE) reporting guideline for cohort studies with the exception of reporting multiple levels of confounder-adjusted estimates (rather than including fully unadjusted estimates).

4 Alimujiang A, Wiensch A, Boss J, *et al.* Association Between Life Purpose and Mortality Among US Adults Older Than 50 Years. *Public Health*; **2**: e194270.
